# The principle of universality and value-based maternity care: a matched population-level study of costs and outcomes for obstetric-led continuity and multiprofessional non continuity models of care

**DOI:** 10.1101/2025.03.04.25323379

**Authors:** Emily J. Callander, Joanne Enticott, Ben W. Mol, Shakila Thangaratinam, Jenny Gamble, Stephen Robson, Helena Teede

## Abstract

**Objective:** We aimed to compare health outcomes and costs in obstetric-led continuity versus multi-professional non-continuity models of care.

**Design:** Observational study with linked administrative data

**Setting:** Australian public and private maternity care

**Population:** 867,334 births, coverall all births in three states of Australia between 2016 and 2019

**Methods:** We analysed outcomes from pregnancy onset to four weeks post-birth in a whole-of-population linked perinatal data asset. Tightly matched cohorts were generated, with bootstrapping of 50 re-matched datasets and sensitivity analyses.

**Main outcome measures:** Stillbirths or neonatal deaths; neonatal intensive care admissions; APGAR score <7 at 5 minutes; 3^rd^ or 4^th^ degree perineal tears; maternal haemorrhages; mean cost per pregnancy episode.

**Results:** Higher adverse outcomes in the multi-professional non-continuity model compared to the obstetric-led continuity model of care, including 786 more stillbirths or neonatal deaths (OR 2.0, 95% CI: 1.8 – 2.1), 2,780 more APGAR score <7 at 5 minutes (OR 2.0, 95% CI: 2.0 – 2.1), 3,327 more 3^rd^ or 4^th^ degree perineal tears (OR 2.9, 95% CI: 2.7 – 3.1) and 10,530 additional maternal haemorrhages (OR 2.7, 95% CI: 2.6 – 2.8). Obesity and mode of birth correlated with neonatal death. Mean cost in AUD per pregnancy episode was $5,888 higher in multi-professional non-continuity model versus obstetric-led continuity, equating to $1.77 billion in extra annual cost to government. Findings persisted across bootstrapping, sensitivity analyses and socioeconomic quintiles.

**Conclusion:** We have shown significant disparity and inequality in outcomes and costs, challenging universal value-based care, with lower adverse health outcomes and costs in the obstetric-led continuity model.

**Funding:** National Health and Medical Research Council (NHMRC).

## Introduction

Advances in the standard of living and maternity care have heralded dramatic improvements in maternal and neonatal health outcomes over the last 100 years, but with significant disparity (1, 2). The recent Lancet Series on Maternity Care highlighted the rapid rise in interrelated, interdependent vulnerabilities, inequitably driving adverse pregnancy outcomes (3). In higher income countries, vulnerabilities include escalating obesity, which align to broader societal trends, driven by eco-social factors, and are concentrated in disadvantaged populations (3). These eco-social factors adversely impact maternal and neonatal outcomes, with devastating impacts on families and increasing health system and economic burden. This highlights the need for adaptive, responsive models of care that deliver value-based healthcare across the domains of *patient and provider experience, quality of care and outcomes, efficiency and sustainability,* considering vulnerabilities, and founded on the tenet of universal healthcare for all (4).

Maternity models of care are complex, with best practice characterised by stakeholder/end-user codesign, evidence-based care with defined outcomes, ongoing monitoring, evaluation and improvement (5). Flawed approaches to care, such as the UK Shrewsbury National Health Service (NHS) Trust, East Kent NHS, and Australian Mackay hospital service failures (6–8), have delivered catastrophic pregnancy outcomes. Independent inquiry all highlighted a failure to investigate, learn and improve. The midwifery continuity model has been compared to standard maternity care in public healthcare settings, (9) with positive outcomes and cost-effectiveness. Yet implementation and scale up remains challenging. There is otherwise a dearth of exploration of broader models of care. In this context, operationalising data in a can explore and optimise models of care across population, profession, continuity and setting. This is key to investigation, learning and improvement of maternity care (20).

Operationalising data in a system that learns and improves is especially relevant to complex maternity models of care in Australia, which includes universal public healthcare supplemented by private care, with embedded mandatory maternity data reporting. Common models include multi-professional, mostly non-continuity care with birth in public hospitals (‘multi-professional, non continuity models of care)’ and obstetric led, continuity of care with birth in private hospitals (‘obstetric-led continuity models of care’). Leveraging our unique linked population level data assets, we aimed to i) quantify differences in value-based care across health outcomes and costs between obstetric-led continuity and multi-professional, non continuity models of care in Australia, comparing tightly matched cohorts by demographic/eco-social and clinical characteristics, including across socioeconomic groups; ii) explore relationships between potential drivers and adverse outcomes.

## Methods

### Study design and participants

This retrospective observational study was conducted across the Australian healthcare system, which aims for equity of access as a fundamental principal, delivered through a dual public and private care system (10), for ∼300,000 births annually. A description of the Australian maternity system is provided in Supplement 1.

#### Data source

The analysis comparing outcomes between births in the obstetric-led continuity and multi-professional, non continuity models of care used the unique Maternity2000 linked data asset, linking routine administrative data on births (covering the pregnancy, birth and postpartum) occurring between January 2016 and December 2019, across the three most populous states in Australia covering 78% of annual births nationally. Women giving birth and their babies were identified from the Perinatal Data Collection, a mandatory dataset of all births, which includes women’s demographic and clinical characteristics prior to and during pregnancy, birth, and postpartum. These data were linked to multiple inpatient, outpatient and emergency department datasets across all private and public hospitals for both the woman and neonate, from onset of pregnancy to 4 weeks post-partum.

The analysis comparing costs between births in the obstetric-led continuity and multi-professional, non continuity models of care, used the unique Maternity1000 data asset (11). This linked data was for all births occurring between January 2016 and June 2018 in one Australian state with 21% of annual births nationally. Women giving birth and their babies were identified from the Perinatal Data Collection, with linkage to the same datasets as Maternity2000, again from onset of pregnancy to 4 weeks post-partum. These data were additionally linked to the MBS and PBS claims records, covering all non-hospital services and prescription pharmaceutical use.

#### Model of care identification

Women who gave birth in a public hospital were classified as being in the ‘multi-professional, non continuity model of care’, as this is the model of care provide in the public hospital system. Women who gave birth in a private hospital were classified as being in the ‘obstetric-led continuity model of care’ as this is the model of care provided in the private hospital system. A full description of each model is provided in Supplement 1.

#### Birth outcomes

Fetal and neonatal outcomes included: stillbirth; neonatal death (death within 28 days of birth); admission to neonatal intensive care unit (NICU); APGAR score <7 at 5 mins; APGAR score <4 at 5 mins; birthweight in 5^th^ centile or less; birthweight in 6^th^ – 10^th^ centile; hypoxic-ischemic encephalopathy (ICD-10-AM code block: P91); birth trauma – including brachial plexus injury, fractured clavicle or humerus or other long bones, peripheral nervous system damage (ICD-10-AM code block: P13.3, P13.4, P14); intrauterine hypoxia (ICD-10-AM code block: P20); other perinatal morbidity – meconium aspiration syndrome, congenital pneumonia or respiratory distress syndrome (ICD-10-AM code block: P24.0, P22, P23). Maternal birth outcomes included: perineal damage – 3rd/4th degree tear; haemorrhage in the woman ICD-10-AM code block: O67, O72), retained placenta (ICD-10-AM code block: O73); and rupture of uterus (ICD-10-AM code block: O71). NICU admission was unavailable for one state.

#### Identification of costs

All health service use covering pregnancy, birth and to 4 weeks postpartum were identified from Maternity1000, along with the corresponding costs, which were summed and presented by different funders: public hospital; MBS; PBS; private health insurers; and individuals through out-of-pocket fees(12). Costs were adjusted for inflation and presented in 2021/22 Australian dollars ($1=0.53 Great British Pounds and 0.67 United States Dollars).

#### Matching populations through strata matching

Matching was used to mimic randomization and to control for key baseline differences in the population of women who gave birth in the obstetric-led continuity and multi-professional, non continuity models of care in both datasets. Matching using simple random sampling without replacement from women who gave birth in the multi-professional, non continuity models of care was conducted based on the number of women who gave birth in a obstetric-led continuity model of care in each stratum group of age, body mass index (BMI), born in a non-English speaking country, socio-economic status, rurality of residence, identification as Aboriginal or Torres Strait Islander, smoking after 20 weeks’ gestation, parity, plurality, Assisted Reproductive Technology (ART) use, and pre-existing diabetes, gestational diabetes, hypertension or preeclampsia. ART and BMI were not available for matching for one state. This produced two equal sized samples with the same distribution of characteristics in the stratum.

### Statistical analysis

Descriptive statistics were completed for demographic and clinical characteristics using the matched cohort from Maternity2000.

Odds ratios of the likelihood of each outcome for multi-professional, non continuity model of care compared to obstetric-led continuity model of care were calculated on matched cohorts. Bootstrapping with 50 rematched datasets was conducted, and the odds ratios produced with each dataset assessed. This bootstrapping generated 50 samples with different matched pairings, ensuring that the findings are not spurious and do not arise from a single matched sample.

Multiple sensitivity analysis were run with the data limited to 1) an intention to treat population, where births at >28 weeks, with women who gave birth in a public hospital with no private obstetric consultations compared to women who initiated care with a private obstetrician prior to 20 weeks’ gestation (any women who were transferred to care in the public system were still classified as being in the private model of care); and 2) a very low risk population including only women between 21 and 35 years, healthy BMI (18.50 – 24.99kg/m^2^), non-smoker after 20 weeks’ gestation, singleton pregnancy, no ART use, and no pre-existing diabetes, gestational diabetes, hypertension or preeclampsia, stratified by nulliparous and multiparous women. The matching was then reconducted and results generated. Further sensitivity analysis stratified outcomes by 1) nulliparous women; 2) multiparous women; 3) nulliparous women giving birth at >=37 weeks’ gestation; 4) multiparous women giving birth at >=37 weeks’ gestation; 5) infants born at different gestational ages; and 6) infants born at different gestational ages with and without congenital abnormalities. Results were also stratified by area-based socioeconomic quintile.

Differences in mean cost between obstetric-led continuity and multi-professional, non continuity models of care in the matched cohorts were analysed. Bootstrapping with 50 rematched datasets was conducted, and the means and difference in means between the two groups produced with each dataset assessed. Three sensitivity analyses were run on the analysis of costs, to assess difference in 1) women who gave birth in a public hospital with no private obstetric consultations compared to women who initiated care with a private obstetrician prior to 20 weeks’ gestation; 2) in a very low risk population (defined above); and 3) with NICU admissions excluded, given its high cost. Matching was then repeated. Results were stratified by area-based socioeconomic quintile.

Descriptive statistics of the number of stillbirths and neonatal deaths that occurred in different birthweight centile groups, gestational age groups, with congenital anomalies, and with an APGAR score of <4 at 5 minutes. An analysis of the likelihood of neonatal death was conducted with multivariable logistic regression, using mode of birth, onset of labour, private model of care, age group, BMI group, born in a non-English speaking country, geographically-based socio-economic status, rurality of residence, identification as Aboriginal or Torres Strait Islander, smoking after 20 weeks’ gestation, parity, plurality, ART use, pre-existing diabetes or gestational diabetes, hypertension and preeclampsia as covariates. This analysis was completed on the full matched cohort, across both model of care. SAS 9.4 was used for all analysis.

## Results

### Outcomes analysis

Overall, there were 661,455 births in multi-professional, non continuity model of care and 202,236 births in obstetric-led continuity model of care in the four-year study period in the three States. Matching produced two equal cohorts of 184,146 women in the obstetric-led continuity and multi-professional, non continuity model of care, with similar characteristics (Table 1).

**Table 1:** Demographic characteristics of women before and after matching, all women giving birth in three states, 2016-2019.

|  | Pre-Matching |  |  |  | Post-Matching |  |  |  |
| --- | --- | --- | --- | --- | --- | --- | --- | --- |
|  | Public model of care<br>(n= 661,455) |  | Private model of care<br>(n=202,236) |  | Public model of care<br>(n=184,146) |  | Private model of care<br>(n=184,146) |  |
|  | *N* | % | *N* | % | *N* | % | *N* | % |
| *Age Group* |  |  |  |  |  |  |  |  |
| Less than 20 years | 23692 | 3.6 | 289 | 0.1 | 225 | 0.1 | 225 | 0.1 |
| 21 - 35 years | 481464 | 73.0 | 122018 | 59.8 | 114741 | 62.3 | 114741 | 62.3 |
| More than 35 years | 154511 | 23.4 | 81717 | 40.1 | 69174 | 37.6 | 69174 | 37.6 |
| Mean age (SD) | 30.4 (5.5) |  | 33.6 (4.3) |  | 32.3 (5.2) |  | 33.4 (4.2) |  |
| *BMI Group^* |  |  |  |  |  |  |  |  |
| Underweight | 16417 | 4.6 | 6489 | 5.4 | 4835 | 4.8 | 4835 | 4.8 |
| Healthy range | 168666 | 46.8 | 67867 | 56.8 | 58233 | 57.5 | 58233 | 57.5 |
| Overweight | 90942 | 25.3 | 27444 | 23.0 | 23420 | 23.1 | 23420 | 23.1 |
| Obese | 84136 | 23.4 | 17601 | 14.7 | 14733 | 14.6 | 14733 | 14.6 |
| Mean BMI (SD) | 34.8 (89.5) |  | 26.6 (41.5) |  | 30.2 (70.3) |  | 26.7 (41.9) |  |
| Mother identifies as Aboriginal or Torres Strait Islander | 36648 | 5.6 | 1002 | 0.5 | 807 | 0.4 | 807 | 0.4 |
| Born in non-English speaking country | 200723 | 30.4 | 53534 | 26.2 | 50361 | 27.4 | 50361 | 27.4 |
| Smoked after 20 weeks' gestation | 59784 | 9.1 | 956 | 0.5 | 717 | 0.4 | 717 | 0.4 |
| Nulliparous | 253231 | 38.4 | 89423 | 43.8 | 81487 | 44.3 | 81487 | 44.3 |
| Singleton pregnancy | 641123 | 97.2 | 197552 | 96.8 | 179369 | 97.4 | 179369 | 97.4 |
| Assisted Reproductive Technology utilised^ | 9683 | 2.7 | 14810 | 12.4 | 7194 | 7.1 | 7194 | 7.1 |
| *Socioeconomic quintile* |  |  |  |  |  |  |  |  |
| 1 (Most disadvantaged) | 146981 | 22.3 | 14263 | 7 | 13498 | 7.3 | 13498 | 7.3 |
| 2 | 146067 | 22.2 | 22432 | 11 | 20281 | 11.0 | 20281 | 11.0 |
| 3 | 146940 | 22.2 | 40060 | 19.7 | 37986 | 20.6 | 37986 | 20.6 |
| 4 | 127159 | 19.3 | 51308 | 25.2 | 47801 | 26.0 | 47801 | 26.0 |
| 5 (least disadvantaged) | 88040 | 13.4 | 74117 | 36.3 | 63686 | 34.6 | 63686 | 34.6 |
| *Rurality of residence* |  |  |  |  |  |  |  |  |
| Major city | 464029 | 70.3 | 173613 | 85.1 | 158688 | 86.2 | 158688 | 86.2 |
| Inner regional area | 123944 | 18.8 | 18260 | 9.0 | 15292 | 8.3 | 15292 | 8.3 |
| Outer regional area | 52236 | 7.9 | 7422 | 3.6 | 6584 | 3.6 | 6584 | 3.6 |
| Rural and remote | 7344 | 1.1 | 953 | 0.5 | 807 | 0.4 | 807 | 0.4 |
| Diabetes (Pre-existing or gestational) | 90033 | 13.65 | 20337 | 9.96 | 18370 | 10.0 | 18370 | 10.0 |
| Hypertension | 18417 | 2.8 | 6371 | 3.1 | 5272 | 2.9 | 5272 | 2.9 |
| Preeclampsia^ | 7553 | 2.1 | 2022 | 1.7 | 1460 | 1.4 | 1460 | 1.4 |

^Not available for one state; BMI= Body Mass Index

In the multi-professional, non continuity model of care, there were 778 more stillbirths or neonatal deaths (0.9% versus 0.4%; OR 2.0 (95% CI: 1.8 – 2.1)), 2,301 more babies admitted to NICU (3.5% versus 1.3%; OR 2.9 (95% CI:2.7 – 3.0)), 2,747 more babies with APGAR score <7 at 5 minutes (3.0% versus 1.5%; OR 2.0 (95% CI: 2.0 – 2.1)) compared to obstetric-led continuity model of care births (Table 2). In the public model of care, there were also 3,373 more women with 3^rd^ or 4^th^ degree perineal tears (2.5% versus 0.7%; OR 2.9 (95% CI: 2.7 – 3.1)), and 10,627 more with haemorrhage (9.6% versus 3.8%; OR 2.7 (95% CI: 2.6 – 2.8)) compared to obstetric-led continuity model of care births. Mode of birth is presented in Table 2, showing lower caesarean sections (31.6% versus 47.9%, p<.0001) and vaginal birth with vacuum (7.2% versus 9.9%, p<.0001), but higher induction of labour (32.5% versus 31.1%, p<.0001) in the obstetric-led continuity model of care. All clinical outcomes show an effect favouring the obstetric-led continuity versus the multi-professional, non continuity model of care (Table 2), with a binomial probability of this occurring by chance of 0·0001 (1 in 8,192).

**Table 2:**
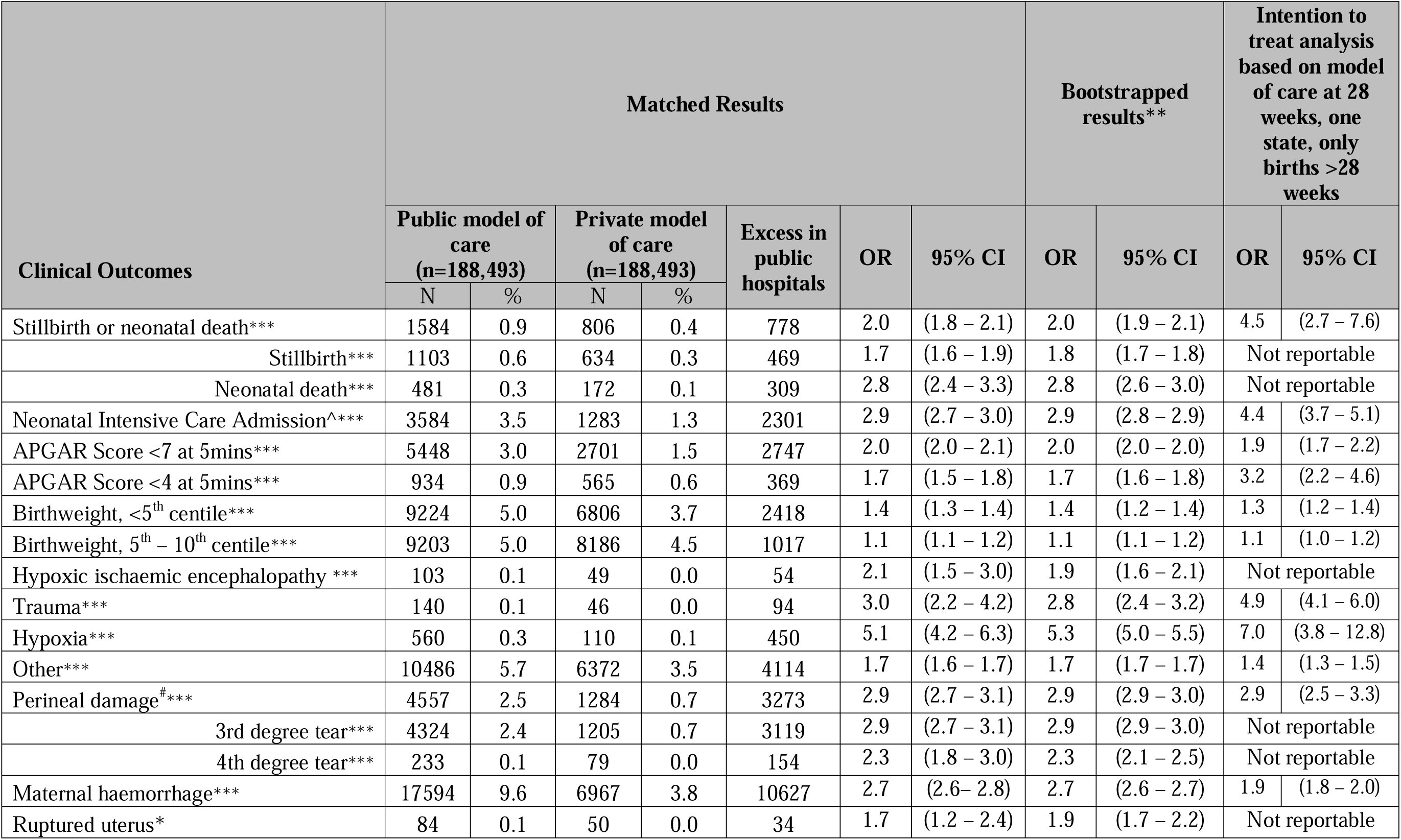
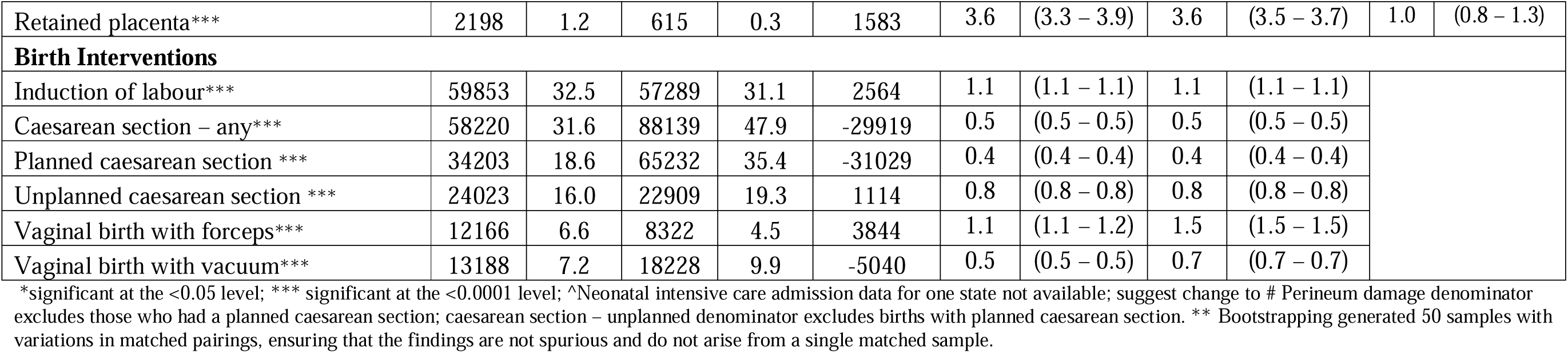
Clinical birth outcomes of women and babies, matched cohort of women giving birth in public and private models of care in three states 2016-2019.

Bootstrapping with 50 rematched samples yielded similar results (Table 2). Sensitivity analysis on intention to treat in the private system was completed in one state with available data. Here, women who initiated obstetric-led continuity and then transferred to the multi-professional, non continuity (public) model were classified in the private model of care, and results were similar to the primary analysis (Table 2). Further sensitivity analysis showed consistent findings across maternal risk and parity, and for infants born over 28 weeks and at 37 weeks gestation and above without congenital abnormalities (Table S1 and S2). Stratifying by socioeconomic quintile, the odds of poor neonatal and maternal outcomes remained consistently higher in the multi-professional, non continuity compared to the obstetric-led continuity model of care (Table S3).

### Cost analysis

Overall, there were 110,041 multi-professional, non continuity model of care births and 38,425 obstetric-led continuity model of care births in the one state with linked available cost data (Table S4). Matching produced two equal-size cohorts of 33,857 women with similar characteristics (Table S4).

Mean costs per birth in AUD for each birth and postpartum episode was $28,645 (95% CI: $28,417 – 28,874) for multi-professional, non continuity model of care and $22,757 (95% CI: $22,624 - $22,890) for obstetric-led continuity model of care (Table 3). Bootstrapped analysis demonstrated that mean costs per pregnancy, birth and postpartum episode was $5,929 (95%CI: $5,789 - $6,081) higher for multi-professional, non continuity model of care compared to obstetric-led continuity model of care (Table 3). Figure 1 shows the difference in average cost to different funders, reflecting the different funding pools for multi-professional, non continuity and obstetric-led continuity model of care. Based upon the annual number of births in Australia (315,507), if the 25% of women currently accessing the obstetric-led continuity model of care were to move to the multi-professional, non continuity model of care, costs to government funders would be $1.77 billion higher per year.

**Table 3:** Mean cost per birth different funders, matched cohort of women giving birth in public and private hospitals in one state, 2016 – 2018.

| Costs | Matched Results |  |  |  | Bootstrapped results** |  |
| --- | --- | --- | --- | --- | --- | --- |
|  | Public model of care<br>(n= 33,857) |  | Private model of care<br>(n=33,857) |  | Difference | 95% CI |
|  | Mean | 95% CI | Mean | 95% CI |  |  |
| Total cost | \$28,645 | (28,417 – 28,847) | \$22,757 | (22,624 – 22,890) | \$5,929 | (5,789 – 6,081) |
| Public hospitals funders | \$26,499 | (26,273 – 26,724) | \$1,739 | (1,629 – 1,849) | \$24,867 | (24,724 – 25,015) |
| Private health insurers | \$111 | (99 – 122) | \$12,921 | (12,862 – 12,981) | -\$12,827 | (-12,838 – -12,816) |
| Medicare | \$1,564 | (1,552 – 1,576) | \$3,812 | (3,791 – 3,832) | -\$2,261 | (-2,268 – -2,255) |
| Pharmaceutical Benefits Scheme | \$99 | (92 – 107) | \$135 | (128 – 143) | -\$36 | (-42 - -28) |
| Patient out-of-pocket | \$472 | (463 – 481) | \$4,285 | (4,266 – 4,304) | -\$3,850 | (-3,856 – -3,845) |

\*\* Bootstrapping generated 50 samples with variations in matched pairings, ensuring that the findings are not spurious and do not arise from a single matched sample.

In the sensitivity analysis with the intention to treat selection of women in the obstetric-led continuity mode of care, mean costs per birth remained comparatively higher for multi-professional, non continuity model of care ($28,391, 95% CI: $20,104 - $31,448 versus $23,432, 95% CI: $18,628 - $25,479). In the sensitivity analysis limited to low-risk women, mean costs per birth remained comparatively higher for multi-professional, non continuity model of care for nulliparous women with similar results found for multiparous women (Table S5). In sensitivity analysis excluding births with a NICU admission, costs remained higher in multi-professional, non continuity model of care (Table S5). Costs also remained consistently higher when stratified by socioeconomic status for women in the multi-professional, non continuity model of care group than the obstetric-led continuity model of care (Table S6).

#### Associations with adverse outcomes

In the overall matched cohort of 376,986 birth across three states, factors associated with stillbirth or neonatal death are identified in Table 4. Vulnerabilities including maternal BMI category, age and maternal birth in a non-English speaking country, were associated with birth outcomes. Mode of delivery was associated with stillbirth or neonatal death, with caesarean section having an adjusted odds ratio (aOR) of 0.3 (0.3 – 0.4) and induction having an aOR of 2.5 (2.2 – 2.8). The obstetric-led continuity model of care had an aOR of 0.7 (0.7 – 0.8) for neonatal death. Higher stillbirths and neonatal deaths in multi-professional, non continuity models than in obstetric-led continuity models, was observed across all birthweight centile and gestational age groups, those with congenital anomalies, and those with an APGAR score of <4 at 5 minutes (Table S7).

**Table 4:**
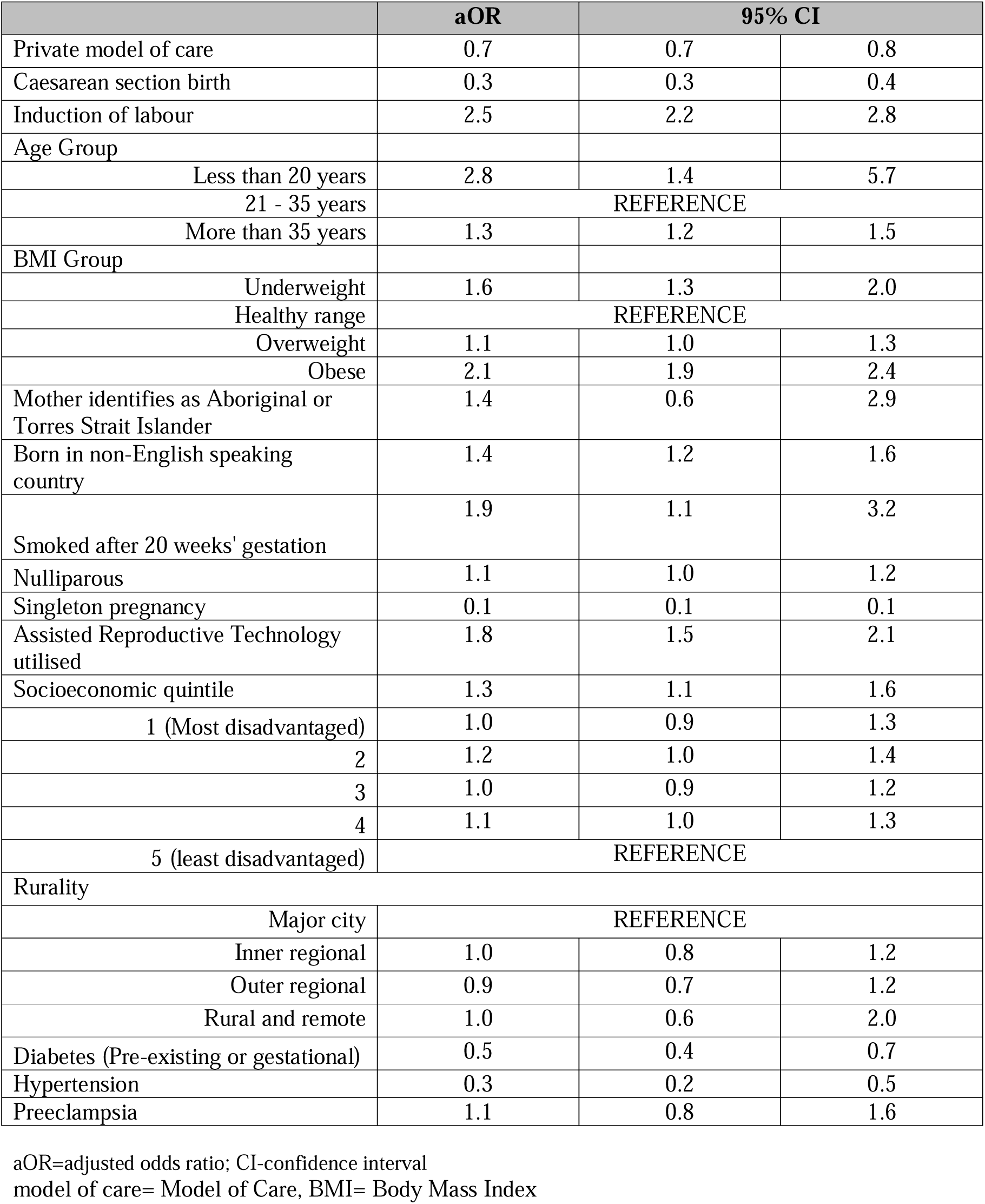
Independent associations with stillbirth or neonatal death in matched cohorts of women giving birth in public and private models of care in three states 2016 - 2019.

|  | aOR | 95% CI |  |
| --- | --- | --- | --- |
| Private model of care | 0.7 | 0.7 | 0.8 |
| Caesarean section birth | 0.3 | 0.3 | 0.4 |
| Induction of labour | 2.5 | 2.2 | 2.8 |
| Age Group |  |  |  |
| Less than 20 years | 2.8 | 1.4 | 5.7 |
| 21 - 35 years | REFERENCE |  |  |
| More than 35 years | 1.3 | 1.2 | 1.5 |
| BMI Group |  |  |  |
| Underweight | 1.6 | 1.3 | 2.0 |
| Healthy range | REFERENCE |  |  |
| Overweight | 1.1 | 1.0 | 1.3 |
| Obese | 2.1 | 1.9 | 2.4 |
| Mother identifies as Aboriginal or Torres Strait Islander | 1.4 | 0.6 | 2.9 |
| Born in non-English speaking country | 1.4 | 1.2 | 1.6 |
| Smoked after 20 weeks' gestation | 1.9 | 1.1 | 3.2 |
| Nulliparous | 1.1 | 1.0 | 1.2 |
| Singleton pregnancy | 0.1 | 0.1 | 0.1 |
| Assisted Reproductive Technology utilised | 1.8 | 1.5 | 2.1 |
| Socioeconomic quintile | 1.3 | 1.1 | 1.6 |
| 1 (Most disadvantaged) | 1.0 | 0.9 | 1.3 |
| 2 | 1.2 | 1.0 | 1.4 |
| 3 | 1.0 | 0.9 | 1.2 |
| 4 | 1.1 | 1.0 | 1.3 |
| 5 (least disadvantaged) | REFERENCE |  |  |
| Rurality |  |  |  |
| Major city | REFERENCE |  |  |
| Inner regional | 1.0 | 0.8 | 1.2 |
| Outer regional | 0.9 | 0.7 | 1.2 |
| Rural and remote | 1.0 | 0.6 | 2.0 |
| Diabetes (Pre-existing or gestational) | 0.5 | 0.4 | 0.7 |
| Hypertension | 0.3 | 0.2 | 0.5 |
| Preeclampsia | 1.1 | 0.8 | 1.6 |

aOR=adjusted odds ratio; CI-confidence interval model of care= Model of Care, BMI= Body Mass Index

## Discussion

### Main findings

Given increasing maternal eco-social vulnerabilities and rising costs of maternity care, we used unique population-level linked administrative data to compare value-based care across outcomes and costs. In a matched Australian population, a multiprofessional, non-continuity model of care (provided in the public hospital system) was compared to the obstetric led, continuity model of care (provided in the private hospital system). All 13 clinical outcomes were better in the obstetric-led continuity and multi-professional, non continuity model of care, with a probability of all being due to chance of 0·0001, supporting true differences. Additional neonatal adverse outcomes in the public model of care included 786 more stillbirths or neonatal deaths, 2,251 NICU admissions, 2,780 APGAR scores <7 at 5 minutes. Additional maternal adverse outcomes in the multi-professional, non continuity model of care included 3,169 with 3^rd^ or 4^th^ degree tears and 10,530 haemorrhages. More induction of labour and less caesarean section occurred in the multi-professional, non continuity model of care. These outcomes were delivered for at least $5,929 per birth more in the multi-professional, non continuity model of care; with additional annual costs of $1.77 billion to government if all women in the obstetric-led continuity model of care were to move to the multi-professional, non continuity model of care. An intention to treat analysis for those who started in obstetric-led continuity care showed consistent results, as did bootstrapping, multiple sensitivity analyses and stratification by socioeconomic status. On the overall matched cohort, maternal vulnerabilities including higher BMI, as well as mode of birth were associated with neonatal death, including an inverse association with caesarean section birth.

### Strengths and Limitations

Limitations include a lack of clarity on the drivers of the observed differential outcomes as current administrative data doesn’t provide insights on the complexity of models of care and costs. We are unable to match for all differences between populations, however measurable parameters suggested well balanced populations. The key strength of this study is the robust propensity matching and the use of administrative data to uniformly capture costs for births in the models of care, including all funding sources, improving the accuracy of reported costs. This allowed capture of all health service use costs throughout pregnancy and birth, aligned to the concept of value-based healthcare across the full healthcare episode, rather than focusing on birth alone(4).

### Interpretation

Differences in outcomes across complex models of care are commonly dismissed or ascribed to population differences such as those in education and socioeconomic status (13). In the current study, leveraging unique datasets, populations were robustly matched to mimic randomization and mitigate population differences. Bootstrapping and multiple sensitivity analyses showed consistent results. Propensity matching cannot capture all differences including social factors (e.g. women’s engagement in care and domestic violence) healthcare provider factors (e.g. health professional training), and health service factors (e.g. NICU access). Likewise, randomized controlled trials can have narrow inclusion criteria, limited relevance to real world populations and challenges in extrapolation across diverse models of care. (14) This mandates both healthcare embedded observational and clinical research focused on improving outcomes.

Modern maternity care has seen dramatic improvements (15), yet our findings mandate research to identify and mitigate driving factors and to optimise models of care across populations, providers, care delivery, settings and resourcing (5). Indeed, service failures are heralded when services fail to lean and improve, noted in the investigating reports from the UK and Australia, calling for embedded, iterative real time research in maternity care (4, 7, 8). The current study accessed the most recent available data (up to 2019), with unchecked inequitable outcomes, lack of transparency and awareness across providers, community and policy makers for five years. This is consistent with Australian maternity data system mapping showing fragmentation, poor harmonisation and linkage, delayed access, unnecessary duplication and bureaucracy and socio-cultural and political barriers to change (16). This is common globally, despite many countries with routine maternity data collection, advanced technology and secure data environments (17). These findings emphasise the need for Clinical Quality Registries for timely, independent, transparent and accessible deidentified health data, shown to underpin improved models of care and outcomes (16). However, accessible data, propensity matching, advanced analytics and benchmarking are not sufficient to improve healthcare in isolation. Learning Health Systems (LHS) incorporating evidence from stakeholders and patients, evidence from research, evidence from practice (data) and evidence from embedded implementation research, comparative effectiveness and economic analyses improve health outcomes(18, 19). Ongoing catastrophic maternity service failures, rising eco-social vulnerabilities, and inequities and disparities such as those demonstrated here, mandate transparent investigation, learning and improvement, that would be enabled by Clinical Registries and LHS approaches.

To our knowledge, this is the first report on value-based differences in matched populations across outcomes and costs, between the multiprofessional, non-continuity model of care and obstetric led, continuity model of care. In unmatched populations, a prior Australian study identified poorer outcomes in public compared to private hospital births (13). Another Australian study compared public hospital midwifery care continuity (best costs and outcomes), multiprofessional (worst costs and outcomes) and obstetric antenatal care, showing APGARs<7 (2.1%, 3.4% and 2.3% respectively) and neonatal death or stillbirth (0.41%, 1.73%, 1.05%) respectively (20). Other studies reported outcomes for non-continuity obstetric or midwifery care without significant differences in outcomes (21–23), potentially indicating the critical role of continuity of care provider in outcomes. Randomised trials on models of care have focused on continuity of midwifery-led model of care, compared to other public models of care and show lower costs and equivalent or better outcomes (9). Consistently, the current study shows comparative lower adverse events in the private obstetric continuity of care model, with potential benefits including clear provider responsibility and strong relational aspects between women and providers, with evidence of better experience (24).

The current study shows that higher BMI and mode of birth were associated with neonatal death. In the context of the continued rise in caesarean section rates, the public health system in Australia, the UK and elsewhere has focused on promoting ‘normal birth’(6, 25). Here, we demonstrate higher caesarean rates in a model of care with better outcomes and costs, alongside inverse correlations between stillbirth or neonatal death, mandating further research on appropriate indications in the context of rising maternal risks. Other key correlates of adverse outcomes included rapidly rising maternal obesity, driven by eco-social factors, highlighting the need for further research and contemporary population data to inform policies on mode of birth.

## Conclusion

Improving equity and value across costs and outcomes is a fundamental priority in maternity care (4, 26). Using unique population level, linked data assets, we have demonstrated significantly worse outcomes for women and neonates, and higher costs, in multi-professional, non continuity compared to obstetric-led continuity obstetric models of care. Potential drivers include differences in continuity of care, rising maternal vulnerabilities and mode of birth. Overall, this study highlights the need for further research, embedded in clinical care. Given the stark disparities and inequality in outcomes between models of care, we propose that a National Maternity Learning Health System are vital to embed implementation and comparative effectiveness research in practice, identify drivers and enable rapid improvement in models of care and outcomes.

## Data Availability

Our data access approvals specifically prohibit any data sharing, and thus data sharing is not possible.

## Author contributions

EC conceived the original study idea and undertook the analysis. EC and HT drafted the manuscript. EC, JE, BM and HT co-designed the analysis. All authors contributed to the interpretation of the findings, and edited the final manuscript.

## Ethics statement

For Maternity1000, ethics approval was obtained from the Townsville Hospital and Health Service Human Research Ethics Committee (HREC) (HREC/16/QTHS/223), James Cook University HREC (H7246) and the Australian Institute of Health and Welfare HREC (EO2017-1-338). We also obtained Public Health Act Approval (RD007377). The Maternity2000 project received human research ethics approval from the New South Wales Health Service Human Research Ethics Committee (HREC) (HREC/ETH00684/2020.11), and the Australian Institute of Health and Welfare Ethics Committee (EO2020/4/1167). We also received Public Health Act Approval (PHA 20-00684). No identifiable patient information was provided to the authors.

## Acknowledgements

The authors acknowledge and thank the Consultative Councils Unit and the Consultative Council on Obstetric and Paediatric Mortality and Morbidity (CCOPMM); the Queensland Statistical Services Branch; the Centre for Health Record Linkage and NSW Ministry of Health for the data and support provided to this project. The opinions and conclusions expressed in this paper are those of the authors and may not reflect those of CCOPMM, Queensland Health, New South Wales Ministry of Health or Victoria Health.

The authors also acknowledge and thank for their support with data extraction.

