## Supplement Files for "The principle of universality and value-based maternity care: a matched population-level study of costs and outcomes for obstetric-led continuity and multiprofessional non continuity models of care" (from another version of this preprint: version 1, 2025-03-05)

**Supplement 1**

Around 75% of births occur in public hospitals (13) with a public model of care generally providing antenatal, intrapartum and postnatal care in a public hospital setting, applying risk stratification, staffed by rostered non-continuity midwives, obstetric registrars and residents with consultant obstetric involvement for those at higher risk. There are policy, funder, consumer, service and provider drivers to lower birth intervention rates in the public system (14, 15). Other providers, including general practitioners may be involved in care alongside allied health professionals as required . Around 25% of births occur in a private hospital with a private model of care that is obstetric led, has patient choice of provider and continuity of care with birth interventions involving shared decision making between the provider and woman and higher rates of caesarean sections. Antenatal and postnatal care following discharge occurs at the private obstetrician’s consulting rooms, and birth occurs in a private hospital with midwifery support .

In health system funding and service management, care outside of public hospitals is largely by private providers with government subsidy through the Medicare Benefits Schedule (MBS), and usually out-of-pocket fees for patients. Pharmaceuticals are government subsidised through the Pharmaceutical Benefits Scheme (PBS), with out-of-pocket fees for some patients. Hospital-based care is provided through public hospitals that are government managed and funded with no out-of-pocket fees; or through private hospitals that are privately managed with some government subsidy through the MBS, but are largely funded through optional private health insurance held by individuals, and out-of-pocket fees.

**Table S1: Sensitivity analysis of neonatal and maternal birth outcomes based upon women’s characteristics, matched cohort of women giving birth in public and private** **models of care** **in three states 2016 – 2019.**

|  | ***Low risk, Nulliparous Women*** | | ***Low risk, Multiparous Women*** | | ***Nulliparous Women*** | | ***Multiparous Women*** | | ***Nulliparous Women, >37 weeks*** | | ***Multiparous Women, >37 weeks*** | |
| --- | --- | --- | --- | --- | --- | --- | --- | --- | --- | --- | --- | --- |
| ***OUTCOMES*** | ***OR*** | ***95% CI*** | ***OR*** | ***95% CI*** | ***OR*** | ***95% CI*** | ***OR*** | ***95% CI*** | ***OR*** | ***95% CI*** | ***OR*** | ***95% CI*** |
| Neonatal death or stillbirth | 2.0 | (1.6 – 2.5) | 2.1 | (1.7 – 2.6) | 2.0 | (1.8 – 2.3) | 1.9 | (1.7 – 2.2) | 2.0 | (1.3 – 3.1) | 2.4 | (1.7 – 3.5) |
| Stillbirth | 1.7 | (1.4 – 2.2) | 2.0 | (1.5 – 2.5) | 1.8 | (1.5 – 2.1) | 1.7 | (1.5 – 2.0) | 1.9 | (1.2 – 3.2) | 2.2 | (1.4 – 3.4) |
| Neonatal death | 3.7 | (2.3 – 6.1) | 2.7 | (1.6 – 4.3) | 3.0 | (2.3 – 2.8) | 2.7 | (2.1 – 3.4) | 2.1 | (0.8 – 5.6) | 2.9 | (1.6 – 5.4) |
| Neonatal Intensive Care Admission^ | 2.9 | (2.4 – 3.5) | 2.5 | (2.1 – 3.0) | 2.9 | (2.6 – 3.2) | 2.8 | (2.6 – 3.1) | 3.8 | (3.0 – 4.8) | 4.4 | (3.6 – 5.3) |
| APGAR Score <7 at 5mins | 1.7 | (1.6 – 1.9) | 2.3 | (2.1 – 2.6) | 1.9 | (1.8 – 2.0) | 2.2 | (2.1 – 2.3) | 2.3 | (2.0 – 2.7) | 2.7 | (2.4 – 3.1) |
| APGAR Score <4 at 5mins | 1.8 | (1.5 – 2.1) | 2.5 | (2.1 – 2.9) | 1.9 | (1.7 – 2.1) | 2.2 | (2.0 – 2.5) | 2.1 | (1.5 – 2.9) | 2.9 | (2.3 – 3.6) |
| Birthweight, <5th centile | 1.4 | (1.3 – 1.4) | 1.4 | (1.2 – 1.5) | 1.4 | (1.3 – 1.5) | 1.3 | (1.3 – 1.4) | 1.8 | (1.7 – 2.0) | 1.9 | (1.8 – 2.1) |
| Birthweight, 5th-10th centile | 1.1 | (1.1 – 1.2) | 1.1 | (1.0 – 1.2) | 1.1 | (1.1 – 1.2) | 1.1 | (1.1 – 1.2) | 1.3 | (1.2 – 1.4) | 1.4 | (1.3 – 1.5) |
| Trauma | 4.8 | (2.0 – 11.5) | 1.7 | (0.7 – 4.3) | 3.5 | (2.3 – 5.6) | 2.5 | (1.5 – 4.1) | 4.3 | (1.8 – 10.1) | 3.1 | (1.2 – 7.8) |
| Hypoxia | 4.6 | (3.2 – 6.8) | 6.5 | (3.6 – 11.6) | 4.2 | (3.3 – 5.4) | 7.3 | (5.0 – 10.4) | 3.5 | (1.9 – 6.6) | 7.3 | (3.5 – 15.0) |
| Other | 1.9 | (1.8 – 2.1) | 1.5 | (1.4 – 1.7) | 1.8 | (1.8 – 1.9) | 1.6 | (1.5 – 1.6) | 1.9 | (1.7 – 2.1) | 1.7 | (1.6 – 1.9) |
| Perineal damage^#^ | 2.8 | (2.6 – 3.0) | 3.3 | (2.9 – 3.7) | 3.5 | (3.2 – 3.7) | 4.2 | (3.7 – 4.7) | 3.7 | (3.2 – 4.4) | 4.9 | (3.8 – 6.5) |
| Maternal haemorrhage | 2.9 | (2.7 – 3.1) | 2.6 | (2.4 – 2.7) | 2.9 | (2.8 – 3.0) | 2.5 | (2.4 – 2.6) | 2.8 | (2.6 – 3.0) | 2.6 | (2.5 – 2.8) |
| Retained placenta | 3.3 | (3.7 – 4.0) | 2.8 | (2.3 – 3.5) | 3.8 | (3.3 – 4.4) | 3.4 | (3.0 – 3.9) | 4.3 | (3.3 – 5.5) | 4.4 | (3.5 – 5.5) |

^Neonatal intensive care admission data for one state not available; # Perineum damage denominator excludes those who had a planned caesarean section; caesarean section – unplanned denominator excludes births with planned caesarean section.

**Table S2: Sensitivity analysis of neonatal birth outcomes based upon neonate’s characteristics, matched cohort of women giving birth in public and private** **models of care in three states, 2016 – 2019**

|  | ***<=28 weeks gestation*** | | ***29 – 32 weeks gestation*** | | ***33 – 36 weeks gestation*** | | ***>=37 weeks gestation*** | | ***<28 weeks gestation with congenital abnormalities*** | | ***>28 weeks gestation with congenital abnormalities*** | | ***<28 weeks gestation without congenital abnormalities*** | | ***29 – 32 weeks gestation without congenital abnormalities*** | | ***33 – 36 weeks gestation without congenital abnormalities*** | | ***>=37 weeks gestation without congenital abnormalities*** | |
| --- | --- | --- | --- | --- | --- | --- | --- | --- | --- | --- | --- | --- | --- | --- | --- | --- | --- | --- | --- | --- |
| ***OUTCOMES*** | ***OR*** | ***95% CI*** | ***OR*** | ***95% CI*** | ***OR*** | ***95% CI*** | ***OR*** | ***95% CI*** | ***OR*** | ***95% CI*** | ***OR*** | ***95% CI*** | ***OR*** | ***95% CI*** | ***OR*** | ***95% CI*** | ***OR*** | ***95% CI*** | ***OR*** | ***95% CI*** |
| Neonatal death or stillbirth | 2.4 | (2.1 – 2.8) | 0.8 | (0.6 – 1.1) | 1.9 | (1.5 – 2.4) | 2.2 | (1.7 – 3.0) | 1.3 | (0.8 – 2.0) | 0.9 | (0.6 – 1.4) | 3.2 | (2.6 – 4.0) | 0.8 | (0.5 – 1.3) | 1.2 | (0.9 – 1.7) | 2.0 | (1.3 – 3.0) |
| Stillbirth | 1.6 | (1.4 – 1.8) | 0.8 | (0.2 – 1.2) | 1.8 | (1.4 – 2.4) | 2.1 | (1.5 – 2.9) | 0.8 | (0.5 – 1.1) | 1.5 | (0.8 – 2.6) | 2.1 | (1.7 – 2.7) | 0.9 | (0.5 – 1.7) | 1.4 | (0.9 – 2.0) | 2.0 | (1.2 – 3.3) |
| Neonatal death | 3.0 | (2.4 – 3.9) | 0.8 | (0.4 – 1.5) | 2.1 | (1.3 – 3.6) | 2.6 | (1.6 – 4.4) | 2.5 | (1.4 – 4.4) | 0.7 | (0.4 – 1.1) | 3.6 | (2.5 – 5.0) | 0.5 | (0.2 – 1.4) | 0.8 | (0.4 – 1.8) | 2.0 | (1.0 – 4.3) |
| Neonatal Intensive Care Admission^ | 3.4 | (2.8 – 4.2) | 2.5 | (1.9 – 3.2) | 2.8 | (2.5 – 3.2) | 4.2 | (3.6 – 4.8) | 0.8 | (0.6 – 1.3) | 1.2 | (1.0 – 1.4) | 5.2 | (4.1 – 6.7) | 2.7 | (2.1 – 3.6) | 2.9 | (2.5 – 3.2) | 4.5 | (3.8 – 5.3) |
| APGAR Score <7 at 5mins | 2.9 | (2.5 – 3.4) | 1.1 | (0.8 – 1.4) | 1.9 | (1.7 – 2.2) | 2.5 | (2.3 – 2.8) | 1.4 | (0.9 – 2.2) | 1.3 | (1.0 – 1.7) | 4.4 | (3.6 – 5.4) | 1.0 | (0.7 – 1.4) | 1.6 | (1.4 – 1.9) | 2.2 | (1.9 – 2.5) |
| APGAR Score <4 at 5mins | 2.1 | (1.8 – 2.4) | 0.9 | (0.7 – 1.3) | 2.2 | (1.8 – 2.7) | 2.5 | (2.1 – 3.1) | 1.2 | (0.8 – 1.8) | 1.2 | (0.8 – 1.9) | 3.0 | (2.4 – 3.7) | 0.9 | (0.5 – 1.6) | 1.6 | (1.2 – 2.2) | 2.1 | (1.5 – 2.9) |
| Birthweight, <5th centile | 1.0 | (0.7 – 1.3) | 1.4 | (0.8 – 2.6) | 1.7 | (1.5 – 2.0) | 1.9 | (1.8 – 2.0) | Not reportable | | 1.0 | (0.8 – 1.2) | 0.9 | (0.5 – 1.7) | 0.6 | (0.3 – 1.3) | 1.6 | (1.3 – 2.0) | 1.6 | (1.5 – 1.8) |
| Birthweight, 5th-10th centile | 0.5 | (0.4 – 0.8) | 1.3 | (0.7 – 2.3) | 1.3 | (1.1 – 1.5) | 1.4 | (1.3 – 1.5) | Not reportable | | 1.1 | (0.9 – 1.4) | 0.4 | (0.2 – 0.8) | 0.8 | (0.4 – 1.7) | 1.2 | (1.0 – 1.4) | 1.3 | (1.2 – 1.4) |
| Trauma | Not reportable | | Not reportable | | 2.0 | (0.6 – 6.8) | 3.8 | (2.0 – 7.1) | Not reportable | | Not reportable | | Not reportable | | Not reportable | | Not reportable | | 4.8 | (2.0 – 11.2) |
| Hypoxia | Not reportable | | Not reportable | | 1.1 | (0.5 – 2.2) | 5.0 | (3.1 – 8.0) | Not reportable | | Not reportable | | Not reportable | | Not reportable | | Not reportable | | 4.6 | (2.6 – 7.9) |
| Other | Not reportable | | Not reportable | | 1.6 | (1.5 – 1.7) | 1.8 | (1.7 – 1.9) | Not reportable | | Not reportable | | Not reportable | | Not reportable | | Not reportable | | 1.6 | (1.5 – 1.8) |

**Table S3: Outcomes from matched cohort of women giving birth in public and private model of care in three states 2016 *–* 2019**

| ***OUTCOMES*** | **Public model of care (n=188,493)** | | **Private model of care (n=188,493)** | | ***OR*** | ***95% CI*** |
| --- | --- | --- | --- | --- | --- | --- |
|  | ***N*** | ***%*** | ***N*** | ***%*** |  |  |
| ***Socioeconomic Quintile 1 (Most disadvantaged)*** | | | | | | |
| Neonatal death or stillbirth | 151 | 1.9 | 68 | 0.5 | 2.3 | (1.7 – 3.1) |
| Neonatal Intensive Care Admission^ | 161 | 2.6 | 105 | 1.7 | 1.4 | (1.1 – 1.8) |
| APGAR Score <7 at 5mins | 455 | 3.3 | 223 | 1.6 | 2.1 | (1.8 – 2.5) |
| Hypoxia | 42 | 0.3 | 21 | 0.2 | 2.9 | (1.5 – 5.6) |
| Other | 775 | 5.5 | 515 | 3.7 | 1.5 | (1.4 – 1.7) |
| Perineal damage^#^ | 332 | 2.4 | 105 | 0.8 | 2.6 | (2.0 – 3.2) |
| Maternal haemorrhage | 1285 | 9.2 | 569 | 4.1 | 2.5 | (2.3 – 3.8) |
| Retained placenta | 90 | 0.6 | 42 | 0.2 | 2.4 | (1.6 – 3.5) |
| ***Socioeconomic Quintile 2*** | | | | | | |
| Neonatal death or stillbirth | 182 | 0.8 | 99 | 0.5 | 1.9 | (1.5 – 2.5) |
| Neonatal Intensive Care Admission^ | 255 | 2.3 | 157 | 0.9 | 2.6 | (2.0 – 3.3) |
| APGAR Score <7 at 5mins | 665 | 3.1 | 366 | 1.7 | 1.9 | (1.7 – 2.2) |
| Hypoxia | 93 | 0.4 | 22 | 0.1 | 4.4 | (2.4 – 8.0) |
| Other | 1138 | 5.2 | 830 | 3.8 | 1.5 | (1.4 – 1.6) |
| Perineal damage^#^ | 542 | 2.5 | 210 | 1.0 | 2.5 | (2.1 – 3.0) |
| Maternal haemorrhage | 2204 | 10.2 | 1006 | 4.6 | 2.4 | (2.2 – 2.6) |
| Retained placenta | 219 | 1.0 | 89 | 0.4 | 3.2 | (2.4 – 4.2) |
| ***Socioeconomic Quintile 3*** | | | | | | |
| Neonatal death or stillbirth | 377 | 1.0 | 179 | 0.5 | 2.2 | (1.8 – 2.6) |
| Neonatal Intensive Care Admission^ | 587 | 2.7 | 247 | 1.2 | 2.5 | (2.2 – 3.0) |
| APGAR Score <7 at 5mins | 1240 | 3.2 | 547 | 1.4 | 2.3 | (2.1 – 2.6) |
| Hypoxia | 123 | 0.3 | 35 | 0.1 | 3.2 | (2.1 – 4.7) |
| Other | 2262 | 5.8 | 1418 | 3.7 | 1.6 | (1.5 – 1.7) |
| Perineal damage^#^ | 1081 | 2.8 | 342 | 0.9 | 2.5 | (2.2 – 2.8) |
| Maternal haemorrhage | 3584 | 9.3 | 1505 | 3.9 | 2.6 | (2.4 – 2.8) |
| Retained placenta | 244 | 0.6 | 140 | 0.4 | 1.8 | (1.4 – 2.2) |
| ***Socioeconomic Quintile 4*** | | | | | | |
| Neonatal death or stillbirth | 382 | 0.8 | 210 | 0.4 | 1.9 | (1.6 – 2.2) |
| Neonatal Intensive Care Admission^ | 1151 | 3.7 | 412 | 1.3 | 3.1 | (2.8 – 3.5) |
| APGAR Score <7 at 5mins | 1344 | 2.8 | 701 | 1.4 | 1.9 | (1.8 – 2.1) |
| Hypoxia | 170 | 0.4 | 32 | 0.1 | 9.0 | (5.5 – 14.9) |
| Other | 2929 | 6.0 | 1882 | 3.9 | 1.6 | (1.5 – 1.7) |
| Perineal damage^#^ | 1260 | 2.6 | 316 | 0.7 | 3.3 | (2.9 – 3.7) |
| Maternal haemorrhage | 4624 | 9.5 | 2021 | 4.1 | 2.5 | (2.4 – 2.6) |
| Retained placenta | 618 | 1.3 | 209 | 0.4 | 3.3 | (2.8 – 3.9) |
| ***Socioeconomic Quintile 5 (Least disadvantaged)*** | | | | | | |
| Neonatal death or stillbirth | 496 | 0.8 | 267 | 0.4 | 1.8 | (1.6 – 2.1) |
| Neonatal Intensive Care Admission^ | 1408 | 4.0 | 548 | 1.6 | 3.0 | (2.7 – 3.3) |
| APGAR Score <7 at 5mins | 1813 | 2.8 | 938 | 1.5 | 2.0 | (1.8 – 2.1) |
| Hypoxia | 219 | 0.3 | 48 | 0.1 | 6.0 | (4.2 – 8.6) |
| Other | 3642 | 5.7 | 2010 | 3.1 | 1.9 | (1.8 – 2.0) |
| Perineal damage^#^ | 1453 | 2.3 | 384 | 0.6 | 3.1 | (2.8 – 3.5) |
| Maternal haemorrhage | 6045 | 9.4 | 2136 | 3.3 | 3.2 | (3.0 – 3.3) |
| Retained placenta | 1088 | 1.7 | 210 | 0.3 | 5.6 | (4.8 – 4.2) |

**Table S4: Demographic characteristics of women before and after matching, all women giving birth in one state, 2016- 2018**

|  | **Pre-Matching** | | | | **Post-Matching** | | | |
| --- | --- | --- | --- | --- | --- | --- | --- | --- |
|  | **Public model of care (n= 110,041)** | | **Private model of care (n=38,425)** | | **Public model of care (n= 33,857)** | | **Private model of care (n=33,857)** | |
|  | **N** | **%** | **N** | **%** | **N** | **%** | **N** | **%** |
| Age group |  |  |  |  |  |  |  |  |
| 20 and under | 3295 | 3.0 | 35 | 0.1 | 30 | 0.1 | 30 | 0.1 |
| 21-34 years | 85124 | 77.4 | 24837 | 64.6 | 23277 | 68.2 | 23277 | 68.2 |
| 35 years and over | 21622 | 19.7 | 13553 | 35.3 | 10825 | 31.7 | 10825 | 31.7 |
| Mean age (SD) | 29.5 (5.7) | | 33.0 (4.4) | | 31.0 (5.5) | | 32.7 (4.2) | |
| BMI Group | | | | | | | | |
| Underweight | 8100 | 7.4 | 1782 | 4.6 | 1569 | 4.6 | 1569 | 4.6 |
| Healthy range | 52562 | 47.8 | 22373 | 58.2 | 19987 | 58.6 | 19987 | 58.6 |
| Overweight | 25185 | 22.9 | 8686 | 22.6 | 7661 | 22.5 | 7661 | 22.5 |
| Obese | 24194 | 22.0 | 5584 | 14.5 | 4915 | 14.4 | 4915 | 14.4 |
| Mean BMI (SD) | 26.0 (6.5) | | 24.8 (5.2) | | 24.8 (5.3) | | 25.1 (5.7) | |
| Born in non-English speaking country | 6852 | 19.5 | 6852 | 17.8 | 6326 | 18.5 | 6326 | 18.5 |
| Aboriginal or Torres Strait Islander | 8576 | 7.8 | 226 | 0.6 | 174 | 0.5 | 174 | 0.5 |
| Socioeconomic quintile | | | | | | | | |
| 1 (Most disadvantaged) | 10720 | 9.7 | 734 | 1.9 | 2332 | 6.9 | 2332 | 6.9 |
| 2 | 13470 | 12.2 | 1739 | 4.5 | 3946 | 11.7 | 3946 | 11.7 |
| 3 | 10048 | 9.1 | 2129 | 5.5 | 8519 | 25.2 | 8519 | 25.2 |
| 4 | 9336 | 8.5 | 2151 | 5.6 | 10461 | 30.9 | 10461 | 30.9 |
| 5 (least disadvantaged) | 11365 | 10.3 | 3800 | 9.9 | 8584 | 25.4 | 8584 | 25.4 |
| Rurality | | | | | | | | |
| Major city | 68541 | 62.3 | 28024 | 72.9 | 24719 | 72.4 | 24719 | 72.4 |
| Inner regional area | 21235 | 19.3 | 5570 | 14.5 | 5031 | 14.7 | 5031 | 14.7 |
| Outer regional area | 16986 | 15 4 | 4195 | 10.9 | 3867 | 11.3 | 3867 | 11.3 |
| Rural and remote | 3200 | 2.9 | 602 | 1.6 | 495 | 1.5 | 495 | 1.5 |
| Smoked after 20 Weeks | 12699 | 11.6 | 189 | 0.5 | 156 | 0.5 | 156 | 0.5 |
| Nulliparous | 31805 | 28.9 | 13655 | 35.5 | 11642 | 34.1 | 11642 | 34.1 |
| Singleton Pregnancy | 107112 | 97 3 | 37060 | 96.5 | 33344 | 97.7 | 33344 | 97.7 |
| Assisted Reproductive Technology utilised | 3253 | 3.0 | 5168 | 13.5 | 2303 | 6.8 | 2303 | 6.8 |
| Diabetes (Pre-existing or gestational) | 15002 | 13.6 | 3398 | 8.8 | 3024 | 8.9 | 3024 | 8.9 |
| Hypertension | 2968 | 2.7 | 1213 | 3.2 | 935 | 2.7 | 935 | 2.7 |
| Preeclampsia | 2681 | 2.4 | 871 | 2.3 | 624 | 1.83 | 624 | 1.8 |

BMI= Body Mass Index

**Table S5: Sensitivity analysis of cost per birth, matched cohort of women giving birth in public and private hospitals in one state, 2016 – 2018, stratified by socioeconomic status**

| **Costs** | **Public model of care** | | **Private model of care** | | |
| --- | --- | --- | --- | --- | --- |
|  | Mean | 95% CI | Mean | 95% CI | |
| *Low risk, Nulliparous* | | | | | |
| Total cost | $26,863 | (26,267 – 27,464) | $21,506 | | (21,211 – 21,802) |
| *Low risk, Multiparous* | | | | | |
| Total cost | $25,986 | (25,559 – 26,413) | $20,944 | | (20,744 – 21,145) |
| *Excluding births with a NICU admission* | | | | | |
| Total cost | $27,306 | (27,148 – 27,464) | $22,492 | | (22,387 – 22,596) |
| *Based on model of care at 28 weeks, one state, only births >28 weeks* |  |  |  | |  |
| Total cost | $28,391 | (20,104 – 31,448) | $23,432 | | (18,628 – 25,479) |

**Table S6: Cost per birth to different funders, matched cohort of women giving birth in public and private hospitals in one state, 2016 – 2018, stratified by socioeconomic status**

| ***Socioeconomic Quintile*** | **Public model of care (n= 33,857)** | | **Private model of care (n=33,857)** | |
| --- | --- | --- | --- | --- |
|  | **Mean** | **95% CI** | **Mean** | **95% CI** |
| Quintile 1 (Most disadvantaged) | $29,437 | (28,314 – 30,559) | $22,525 | (22,035 – 23,015) |
| Quintile 2 | $29,300 | (28,635 – 29,966) | $22,465 | (22,154 – 22,775) |
| Quintile 3 | $29,583 | (29,072 – 30,095) | $22,759 | (22,528 – 22,990) |
| Quintile 4 | $28,280 | (27,886 – 28,674) | $23,176 | (22,862 – 23,490) |
| Quintile 5 (Least disadvantaged) | $27,659 | (27,288 – 28,030) | $22,452 | (22,260 – 22,643) |

**Table S7: Number of stillbirths and neonatal deaths that occurred in different birthweight centile groups, gestational age groups, those with congenital anomalies, and those with an APGAR score of <4 at 5 minutes, in the matched cohort of women giving birth in public and private hospitals in three states, 2016 – 2019**

|  | **Stillbirth and Neonatal Death** | | **Stillbirth** | | **Neonatal Death** | |
| --- | --- | --- | --- | --- | --- | --- |
|  | *Public* | *Private* | *Public* | *Private* | *Public* | *Private* |
| *Gestational age birthweight centile* | | | | | | |
| <=5th centile | 200 | 73 | 146 | 59 | 54 | 14 |
| 6th-10th centile | 62 | 41 | 49 | 35 | 13 | 6 |
| 11th-25th centile | 204 | 93 | 163 | 80 | 41 | 13 |
| >25^th^ centile | 1118 | 599 | 745 | 460 | 373 | 139 |
| *Gestational age at birth* |  |  |  |  |  |  |
| <=28 weeks | 1030 | 494 | 696 | 397 | 334 | 97 |
| 29 - 32 weeks | 126 | 58 | 99 | 45 | 27 | 13 |
| 33 - 36 weeks | 197 | 119 | 156 | 97 | 41 | 22 |
| >=37 weeks | 231 | 135 | 152 | 95 | 79 | 40 |
| *Congenital Abnormalities* |  |  |  |  |  |  |
| Yes | 567 | 364 | 385 | 268 | 182 | 96 |
| No | 274 | 162 | 181 | 120 | 93 | 42 |
| *APGAR <4, 5 mins* |  |  |  |  |  |  |
| Yes | 115 | 71 | Not applicable | | 114 | 71 |
| No | 726 | 455 |  |  | 161 | 67 |
